# Diaper need is associated with risk for food insecurity in a statewide sample of WIC participants

**DOI:** 10.1101/2020.07.10.20150599

**Authors:** Emily H. Belarmino, Amy Malinowski, Karen Flynn

## Abstract

Diaper need is a form of material hardship that acutely affects families with young children, is not currently addressed by US antipoverty programs, and has received little public or scientific attention. This study examined the association between diaper need and risk for food insecurity in a statewide sample of participants in the Special Supplemental Nutrition Program for Women, Infants, and Children (WIC). Families enrolled in Vermont WIC in August 2019 were invited to an online survey. Generalized linear models were used to estimate the relationship between risk for food insecurity measured by the Hunger Vital Sign tool and diaper need, with and without adjustment for household factors. Follow-up questions asked those with diaper need what they do when they run out of diapers and those without diaper need how they access enough diapers. Complete data were available for 501 households. Over half (52.3%) were at risk for food insecurity and nearly one-third (32.5%) reported diaper need. The odds of experiencing risk for food insecurity were 3.852 (95% CI=2.557, 5.803) times greater for families with diaper need than for families that had enough diapers. The association persisted with adjustment for location, age of respondent, number of children in diapers, and length of time participating in WIC (adjusted OR=4.036, 95% CI=2.645, 6.160). Strategies to avoid running out of diapers included borrowing, stretching supplies, switching to cloth or underwear, and buying on credit. It is possible that public health interventions that address diaper need may reduce food insecurity in households with children.

## 1. INTRODUCTION^1^

In 2018, 14.3% of American households with children aged 0-5 years were food insecure at some point during the previous year, and in 6.7% of those households, children experienced food insecurity as well as adults (Coleman-Jensen et al., 2019). The number of people experiencing food insecurity is projected to increase substantially as closures and social distancing meant to control the spread COVID-19 in 2020 lead to a rise in unemployment and poverty, a known risk factor for food insecurity (Hake et al., 2020; Wight et al., 2014). Food insecurity during early childhood has been associated with poorer general health, developmental risk, and hospitalizations (Drennen et al., 2019), and may be particularly deleterious given the long-term consequences of failing to optimize growth and brain development during early life (Cusick and Georgieff, 2016).

Among the most important federal programs aimed at reducing the effects of poverty on young children are those that provide food benefits, especially the Special Supplemental Nutrition Program for Women, Infants, and Children (WIC) and the Supplemental Nutrition Assistance Program (SNAP; formerly Food Stamps) (Pac et al., 2017). Research suggests that participation in these programs alleviates food insecurity (Kreider et al., 2016; Shaefer and Gutierrez, 2013) and that WIC participation improves diet quality (Tester et al., 2016). Although necessary for all young children, diapers are not an allowable expense in these antipoverty programs nor are they a target of other federal programs such as the Temporary Assistance Program for Needy Families (TANF).

A sufficient supply of diapers is essential for child health and for families living in poverty, the cost of diapering a child can be a source of considerable financial and emotional strain. Failure to provide enough diapers increases risk for diaper dermatitis (Scheinfeld, 2005) and urinary tract infections (Sugimura et al., 2009), as well as parenting stress, a mediator between poverty and child health outcomes (Smith et al., 2013). Diaper need refers to uncertain and insufficient diaper availability and access, and the worry and adverse health outcomes that may result (Smith et al., 2013).

Prior research suggests that food and diaper needs may overlap for many poor families with young children (Austin and Smith, 2017; Massengale et al., 2017). However, these studies did not utilize valid measures of food need. The aims of this study are to quantify diaper need in a statewide sample of low-income families participating in the WIC program and examine the association between diaper need and risk for food insecurity. There is value in understanding links between diaper and food needs, because although these forms of material hardship have the potential to exacerbate one another they are modifiable.

## 2. MATERIALS AND METHODS

### 2.1. Study population

Data were from a cross-sectional survey of Vermont WIC participants. In August 2019, the head of household in all 6905 families participating in the state’s WIC program that accept texts from WIC (93% of all WIC households) were sent a link to the annual online participant survey. A reminder text was sent after one week. This study was determined exempt by the University of Vermont Institutional Review Board (IRB) and Vermont Agency of Human Services IRB.

### 2.2. Measures

The survey asked about WIC implementation, food insecurity, and diaper need. Risk for food insecurity was assessed with the Hunger Vital Sign, a validated, two question screening tool based on the Household Food Security Scale (Hager et al., 2010).

Respondents were asked how many children in their household wear diapers, and those who indicated one or more children in diapers were asked about diaper need. Following Smith et al. (2013), diaper need was assessed with the question, “Do you ever feel that you do not have enough diapers to change them as often as you would like?” Those who responded “yes” were considered to report diaper need and those who responded “no” were considered to report no diaper need. Follow-up questions asked those who reported diaper need what they do when they do not have enough diapers and those who did not report diaper need what they do to have enough diapers.

Demographic factors captured through the survey included age of respondent, length of participation in WIC, and town of residence. We categorized towns in the Burlington-South Burlington Metropolitan New England City and Town Area (NECTA) as metropolitan; all other towns were categorized as non-metropolitan (US Census Bureau, n.d.-a).

### 2.3. Statistical analysis

We used descriptive statistics to summarize the distributions of study variables, and reviewed responses for “other” strategies to access diapers. We used unadjusted generalized linear models with a binary distribution and a logit link to examine the associations between diaper need and each variable of interest. To further examine the association between diaper need and for food insecurity, we used an adjusted model which accounted for demographic variables that had theoretical plausibility: age of respondent, length of participation in WIC, metropolitan/non-metropolitan residence, and the number of children in the household in diapers. Significance was set at α=0.05. All analyses were performed in 2020 using IBM SPSS Statistics version 26 (IBM Corp., Armonk, NY).

Of the 761 individuals who started the survey, we excluded 73 who reported no children in the household in diapers and 187 who were missing data for diaper need or risk for food insecurity. Due to small amounts of missing data for demographic factors, the sample sizes vary slightly between models.

## 3. RESULTS

Consistent with the largely rural composition of Vermont, over two thirds of respondents (72.1%, n=361) lived in non-metropolitan areas (Table 1). Most were over 30 (59.3%, n=297) and had one or two children in diapers (95.6%, n=479).

**TABLE 1.**
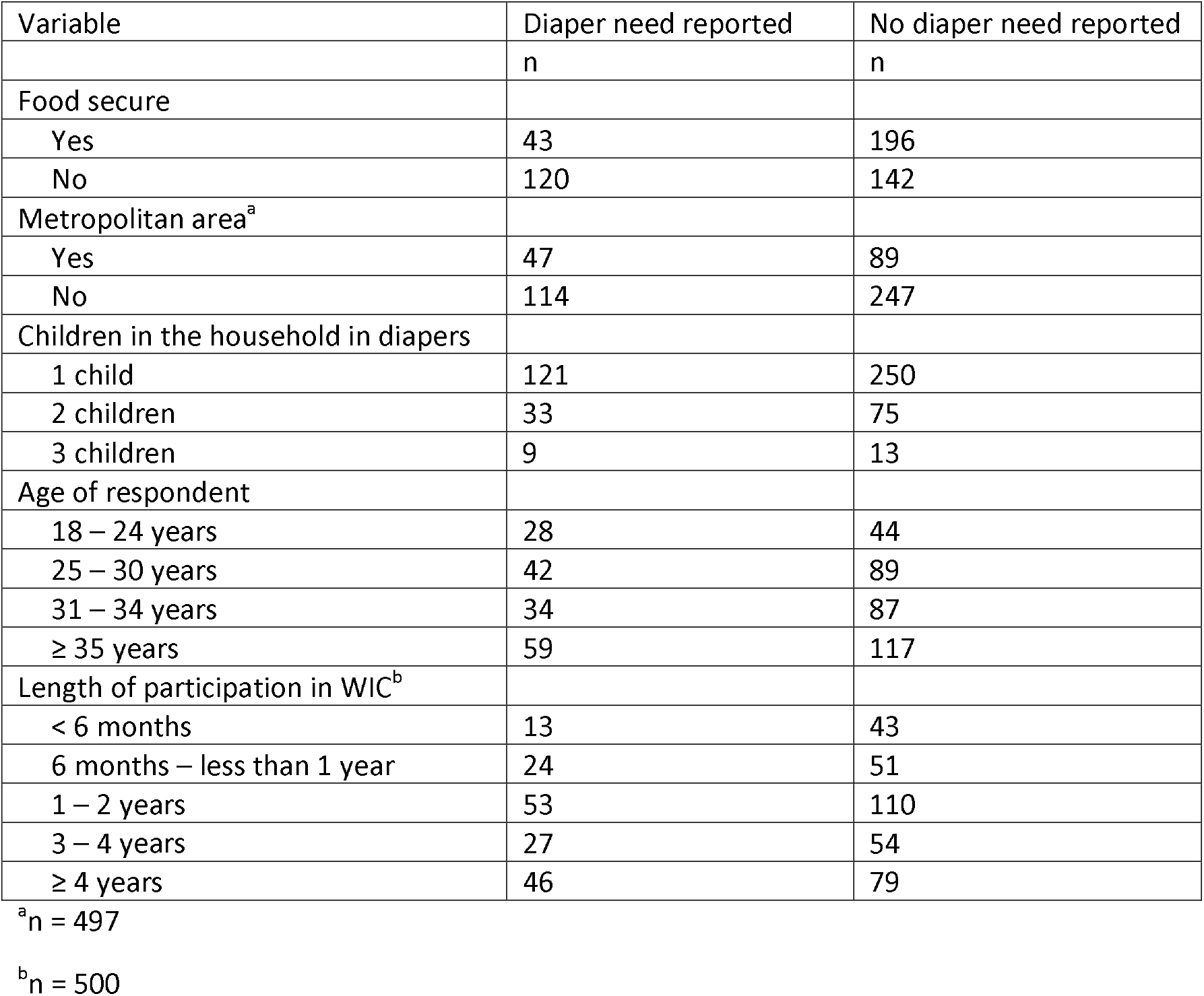
Characteristics of Vermont WIC families with at least one child in diapers (n=501), August 2019

Over 30% of respondents (32.5%, n=161) reported diaper need. Just over half (52.3%, n=258) were at risk for food insecurity. The odds of experiencing risk for food insecurity were 3.852 (95% CI=2.557, 5.803) times greater for families that did not have enough diapers than for families that had enough diapers (Table 2). The association between diaper need and risk for food insecurity persisted with adjustment for demographic factors (AOR=4.036, 95% CI=2.645, 6.160). In the unadjusted and adjusted models, respondents age 31-34 years had higher odds of food insecurity compared to those age 35 years and older (OR=1.646, 95% CI=1.033, 2.624; AOR=1.781, 95% CI=1.083, 2.929).

**TABLE 2.** Predictors of food insecurity among Vermont WIC families with at least one child in diapers (n=496), August 2019

| Variable | Unadjusted OR (95% CI) | Adjusted OR (95% CI) |
| --- | --- | --- |
| **Diaper need** |  |  |
| Yes | 3.852 (2.557, 5.803)** | 4.036 (2.645, 6.160)** |
| No (reference) | 1.00 | 1.00 |
| **Metropolitan area** |  |  |
| Yes (reference) | 1.00 | 1.00 |
| No | 0.845 (0.569, 1.256) | 0.861 (0.565, 1.314) |
| **Children in the household in diapers** |  |  |
| 1 child | 2.115 (0.866, 5.162) | 2.364 (0.869, 6.433) |
| 2 children | 1.566 (0.607, 4.038) | 1.808 (0.632, 5.172) |
| 3 children (reference) | 1.00 | 1.00 |
| **Age of respondent** |  |  |
| 18 – 24 years | 1.641 (0.944, 2.851) | 1.479 (0.802, 2.724) |
| 25 – 30 years | 1.561 (0.991, 2.460) | 1.538 (0.939, 2.519) |
| 31 – 34 years | 1.646 (1.033, 2.624)* | 1.781 (1.083, 2.929)* |
| ≥ 35 years (reference) | 1.00 | 1.00 |
| **Length of participation in WIC** |  |  |
| < 6 months | 0.916 (0.488, 1.721) | 1.043 (0.526, 2.071) |
| 6 months – less than 1 year | 1.322 (0.743, 2.354) | 1.254 (0.662, 2.376) |
| 1 – 2 years | 1.072 (0.673, 1.709) | 1.114 (0.673, 1.842) |
| 3 – 4 years | 1.114 (0.636, 1.949) | 1.188 (0.651, 2.167) |
| ≥ 4 years (reference) | 1.00 | 1.00 |

\* p<0.05, \*\* p<0.001

Of respondents who reported diaper need, 49.1% (n = 80) said that they borrow diapers or money from friends or family, 16% (n = 26) receive diapers from an agency or support organization, 60.7% (n = 99) stretch the diapers that they have when their supply is running short, and 16.6% (n = 27) use other strategies to cope. Of those who reported an adequate supply of diapers, 12.1% (n = 41) said that they borrow diapers or money from friends or family, 5% (n = 17) receive diapers from an agency or support organization, 13.9% (n = 47) stretch the diapers that they have, 87.6% (n = 296) purchase diapers with their own money, and 10.1% (n = 34) use some other strategies. Respondents who reported other strategies often described using cloth diapers or underwear. Some described reallocating their budget, for example, “[I] go into credit card debt for them” or “Money gets taken from food budget to buy diapers.”

## 4. DISCUSSION

To our knowledge, this study is the first to quantify diaper need in a predominantly rural sample and among the first examine the association between diaper and food needs. We found a third of Vermont WIC households with a child in diapers to report diaper need and over half to be at risk of food insecurity. Disposable diapers represented a substantial financial burden for families, and those with diaper need were significantly and substantially more likely to be at risk for food insecurity.

Prior peer-reviewed studies have quantified diaper need among low-income, urban mothers in New Haven, Connecticut using the same survey question as the present study. The New Haven research found 27.5% of mothers with a child under 18 years and 50.3% of mothers with a child aged three years or under in diapers to report diaper need (Austin and Smith, 2017; Smith et al., 2013). The higher levels of diaper need documented in New Haven may be linked to racial disparities. Nearly 95% of Vermont’s population identify as white (US Census Bureau, n.d.-b) compared to 9-15% of the New Haven participants (Austin and Smith, 2017; Smith et al., 2013). A survey of families utilizing a North Carolina diaper bank found a greater percentage to self-identify with a racial or ethnic minority group or speak a language other than English at home compared to the local population (Massengale et al., 2017).

Predictably, the prevalence of risk for food insecurity among our sample was higher than prevalence of food insecurity previously documented among WIC participants using the US Department of Agriculture’s 18-item Household Food Security Survey (HFSS) (Kreider et al., 2016). The Hunger Vital Sign is designed to screen for risk for food insecurity (Hager et al., 2010). Prior testing against the HFSS found the Hunger Vital Sign to have a specificity of 83% indicating that 17% of families who were food secure according to the HFSS were found to be at risk for food insecurity by the Hunger Vital Sign (Hager et al., 2010). Thus, this tool may be effective in identifying families who are vulnerable to future food insecurity as well as families who are already food insecure.

The finding that diaper need is significantly associated with risk for food insecurity has policy relevance and supports the proposition that an adequate supply of diapers may reduce food insecurity. There is potential for WIC staff and pediatric providers to screen for diaper need and food insecurity and refer families to local services. WIC participants are often eligible for other government assistance programs (e.g. nearly all WIC participants are eligible for SNAP, but only one third of Vermont WIC participants indicate SNAP enrollment) and some may benefit from referrals to local food pantries or diaper banks. Future research should explore awareness, perceptions, and use of food pantries among families with diaper need, and the role of food banks in mitigating diaper need on top of growing food needs (Hake et al., 2020). Policy interventions to remove sales tax on diapers or provide a diaper stipend to low-income families also could reduce diaper need. As of January 2020, 36 states charge sales tax on diapers and California is the only state to provide financial assistance for diapers through an antipoverty program (TANF) (National Diaper Bank Network, n.d.).

In this study, examination of demographic variables with possible association to diaper need was limited to those included in the annual WIC participant survey. Questions about diaper need and food security appeared at the end of the survey, which likely impacted the number of responses to these questions. However, reach and internal validity may have been strengthened by the use of an existing statewide survey administered by the WIC program. Although the response rate was similar to the 2018 WIC participant survey, response rates tend to be lower for online surveys compared to other survey formats and challenges reaching disadvantaged populations have been documented (Daikeler et al., 2019; Xu et al., 2019). Large, diverse samples will be required to explore associations between diaper need and a broader range of demographic characteristics. The association between diaper need and risk of food insecurity found in this study needs to be replicated. Cross-sectional data limits the ability to make causal assumptions about observed associations. Future research should consider how interventions to alleviate diaper need impact food security and child health.

## 5. CONCLUSIONS

Although research suggests that nutrition assistance programs reduce food insecurity and other forms of material hardship (Kreider et al., 2016; Shaefer and Gutierrez, 2013), this study calls attention to the fact that many families participating in WIC still struggle to access food and diapers and that these needs may aggravate one another, potentially leading to negative impacts on child health. Interventions to reduce diaper need may mitigate some aspects of child poverty and contribute to improvements in child nutrition and health.

## Data Availability

Data are property of the Vermont Agency of Human Services

## ACKNOWLEDGEMENTS

The authors would like to thank Stephen Parry for statistical support.

## Footnotes

1 IRB: Institutional Review Board; NECTA: New England City and Town Area; SNAP: Supplemental Nutrition Assistance Program; TANF: Temporary Assistance Program for Needy Families; WIC: Special Supplemental Nutrition Program for Women, Infants, and Children

